# Diagnostics for detection and surveillance of priority epidemic-prone diseases in Africa: an assessment of testing capacity and laboratory strengthening needs

**DOI:** 10.1101/2024.05.17.24307542

**Authors:** Aytenew Ashenafi, Olajumoke Sule, Trevor Peter, Silver Mashate, Osborn Otieno, Yenew Kebede, John Oio, Kekeletso Kao, Jane Carter, Toni Whistler, Nqobile Ndlovu, Yenew Kebede

**Affiliations:** Centre of Laboratory Systems Division, Africa Centres for Disease Control and Prevention, Addis Ababa, Ethiopia; International Health Regulations Strengthening Project, Health Protection Operations, United Kingdom Health Security Agency, London, United Kingdom; Clinton Health Access Initiative, Boston, Massachusetts, United States of America; African Society for Laboratory Medicine, Addis Ababa, Ethiopia; Technical Advice and Partnerships Department, The Global Fund to Fight AIDS, Tuberculosis and Malaria, Geneva, Switzerland; Diagnostic System Strengthening Unit, FIND, Geneva, Switzerland; Clinical and Diagnostics Programme, Amref Health Africa, Nairobi, Kenya

## Abstract

In 2023, Africa experienced 180 public health emergencies, of which 90% were infectious diseases and 75% were related to zoonotic diseases. Testing capacity for epidemic-prone diseases is essential to enable rapid and accurate identification of causative agents, and for action to prevent disease spread. Moreover, testing is pivotal in monitoring disease transmission, evaluating public health interventions, and informing targeted resource allocation during outbreaks. An online, self-assessment survey was conducted in African Union Member States to identify major challenges in testing for epidemic-prone diseases. The survey assessed current capacity for diagnosing priority epidemic-prone diseases at different laboratory levels. It explored challenges in establishing and maintaining testing capacity to improve outbreak response and mitigate public health impact. Survey data analysed diagnostic capacity for priority infectious diseases, diagnostic technologies in use, existing surveillance programmes and challenges limiting diagnostic capacity, by country. The survey result from 15 Member States who responded to the survey, showed high variability in testing capacity and technologies across countries and diverse factors limiting testing capacity for certain priority diseases like dengue and Crimean-Congo haemorrhagic fever. At the same time, there is better diagnostic capacity for coronavirus disease 2019 (COVID-19), polio, and measles due to previous investments. Unfortunately, many countries are not utilising multiplex testing, despite its potential to improve diagnostic access. The challenges of limited laboratory capacity for testing future outbreaks are indeed significant. Recent disease outbreaks in Africa have underscored the urgent need to strengthen diagnostic capacity and introduce cost-effective technologies. Small sample sizes and differing disease prioritisation within each country limited the analysis. These findings suggest the benefits of evaluating laboratory testing capacity for epidemic-prone diseases and highlight the importance of effectively addressing challenges to detect diseases and prevent future pandemics.

## Introduction

In 2023, Africa has documented more than 168 public health events affecting 45 African Union Member States mostly infectious disease outbreaks including mpox, diphtheria, dengue, Lassa fever, measles, polio, Rift Valley fever and cholera. These outbreaks continue to expand rapidly and affect many countries on the continent. Since the beginning of 2024, a total of 26,122 cholera cases (3,396 confirmed; 22,726 suspected) with 663 deaths were reported from 12 African countries; 4,451 cases of measles (178 confirmed; 4,273 suspected) and 51 deaths were reported from seven African countries; and for diphtheria 1,573 cases were reported (904 confirmed; 669 suspected) with four deaths from two African countries (1–4).

The International Health Regulations (IHR, 2005) identify disease mapping, health risk assessment and resource prioritisation as among the core capabilities of public health emergency preparedness and response systems (5). Recurrent outbreaks of emerging and re-emerging pathogens have revealed major gaps in these systems in countries on the African continent. Key mechanisms to prevent recurrent disease outbreaks include early detection of infectious diseases in the community, rapid pathogen identification, and integrated disease surveillance. Surveillance and response capacity is limited in many African countries, especially in the context of COVID-19, Ebola virus disease (EVD), and in detection and monitoring of antimicrobial resistance (6). As the recent EVD, COVID-19 and mpox outbreaks highlight, a major paradigm shift is required to establish effective infrastructure and mechanisms for outbreak detection, and common frameworks for preparedness and national public health responses to manage future epidemics (7).

The Africa Centres for Disease Control and Prevention (Africa CDC) has drawn up a preliminary list of priority epidemic-prone diseases for Africa using a risk ranking and analysis tool to identify the target infectious diseases for epidemic preparedness and response actions (8), and to inform research and technology development and disease control innovations such as development and provision of vaccines, diagnostics, and therapeutics. For example, EVD, cholera and COVID-19 scored the highest for disease severity, risk and epidemic potential, as well as the need for preparedness through vaccine availability, and medical and non-medical countermeasures. The disease ranking processes changes every time as more evidence comes in.

The ability to respond effectively to disease outbreaks depends on several factors, including laboratory capacity, a trained health workforce, and robust surveillance systems (9), which are public health systems that rapidly diagnose and contain the spread of infectious diseases. While significant diagnostic capacity has been established over the past two decades for major endemic diseases such as HIV/AIDS, tuberculosis (TB), malaria and more recently COVID-19, testing capacity for other epidemic-prone infections is more limited. Therefore, a comprehensive approach is needed to address the gaps in pandemic preparedness and health systems’ resilience (10, 11).

The Africa CDC undertook a survey of African Union Member States on disease priorities, laboratory capacities and public surveillance systems to help identify major challenges faced in building testing capacity for outbreak detection and response for epidemic-prone diseases in African countries. The findings of this survey are presented in this paper.

## Methods

### Survey description

All 55 African Union Member States were invited to participate in a self-assessment survey of laboratory testing capacity and surveillance of 22 epidemic-prone diseases between February and April 2023. These diseases were selected through a risk ranking and prioritisation exercise undertaken by Africa CDC in collaboration with the European Centre for Disease Prevention and Control in 2022 (8).

### Survey instrument

The survey instrument was developed and validated in consultation with members of the Africa Laboratory Technical Working Group (S1 questionnaire file). It included questions on disease testing capacity, technology in use, healthcare system level and presence of surveillance programmes. Additionally, questions addressed multiplex testing capacity and challenges faced.

Participants were asked to rank their capacity to diagnose the priority epidemic-prone diseases (S2 list file) on a five-point Likert scale (from 1–5), with 5 being the highest. Quantitative questions included information on disease testing capacity across countries’ healthcare systems, standard methods used to diagnose diseases (PCR, multiplex PCR, lateral flow assays, ELISA, etc.), biosecurity, sample transport, data systems, staffing and existing surveillance programmes. Qualitative questions were also included to allow respondents to add relevant comments, and to identify countries’ most significant challenges and the support needed to ensure adequate diagnostic capacity for future outbreaks and epidemics. The survey tool was developed in a Microsoft Excel format using dropdown lists to make it user-friendly. The tool was provided in English and French, pre-tested, amended and distributed online to respondents (S1 questionnaire file).

The first section of the survey comprised 8-questionnaire for disease testing capacity to correlate the epidemic-prone disease to the capacity of diagnosis and technology in use and surveillance programmes for these diseases. The respondent only completes the boxes. Some of these boxes have a dropdown menu so that they can only select answers. The tool allows countries to add a comment describing their testing method and asking about the standard methods used to diagnose diseases (PCR, Multiplex PCR, Lateral Flow Assay, ELISA, and others). Based on the responses, the ability to diagnose the twenty-two priority epidemic-prone diseases were rated on a five-point scale from 1 to 5, with 5 representing the highest score..

The second survey has 3-questionnaire for multiplex testing capacity and panel of multiplex testing, and levels of laboratory testing capacity across Member States’ healthcare systems.

The third survey has 8-miscellaneous and 3-qualitative questionnaires on sample transport, biosecurity, data systems and staffing. Some qualitative questions were included to get a general sense of the support countries need/want and explore countries’ biggest challenges to ensuring diagnostic capacity to be ready for a future epidemic.

### Dissemination of the survey tool to countries and data collection

All African countries were invited to participate in the survey, targeting national public health laboratory directors, managers and heads of reference laboratories. We collected the informed consent form from online survey links obtained from a network of laboratories that got a request to participate in a survey. Data were collected from February to April 2023.

### Data entry and analysis

Data were entered into SPSS 2023 and Excel for data analysis and visualisation.

### Ethics Considerations

Ethics approval was not sought for the following reasons.

The survey pertained to member state diagnostic capacity did not include data on individual patients or vulnerable people. The survey instrument was developed in consultation with members of Africa CDC Laboratory Technical Working Group. Participation by member states was voluntary following informed consent. Responses were stored securely on Africa CDC platform and published results of the survey Responses were via online methodology and results of the survey are anonymised and cannot be traced to member states.

## Results

### Survey respondents

Participants from 15 of 55 (27%) Member States responded to the survey (Fig 1). Respondents were distributed as follows: five (50%) from the southern region, four (27%) from the western region, three (21%) from the eastern region, two (29%) from the northern region, and one (11%) from the central region. The survey was completed by laboratory directors (7) and senior technical staff members (8) working at either national public health institutes (12) or ministries of health (3).

**Fig 1.**
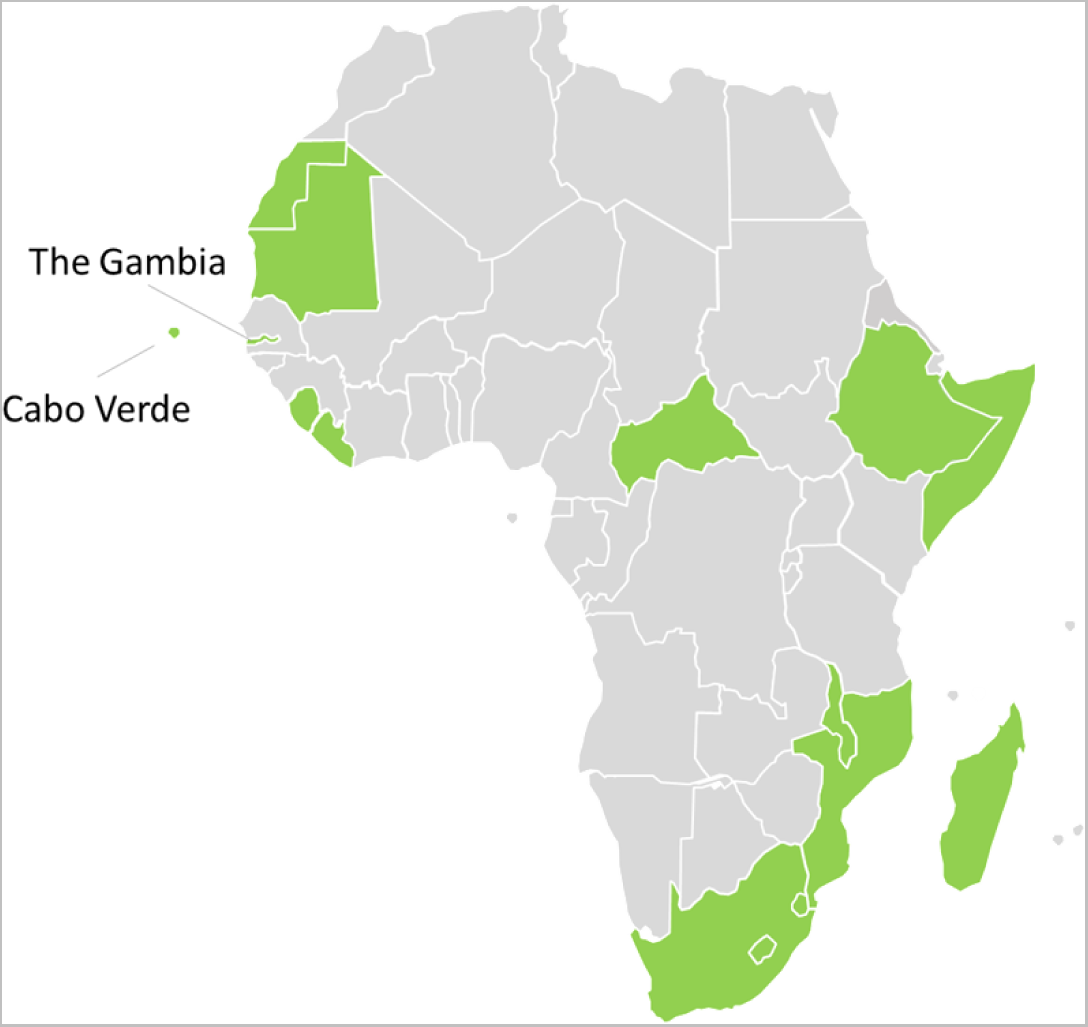
Map showing countries that participated in the survey (n=15)

### Disease testing capacity by laboratory level

Capacity for diagnosing infectious diseases varied based on the disease, country and laboratory level (Fig 2). Testing for viral haemorrhagic fevers was conducted at central public health laboratories and tertiary care hospital laboratories, while testing for COVID-19, meningococcal meningitis, cholera, measles and dengue was conducted at laboratories ranging from reference to district level.

**Fig 2.**
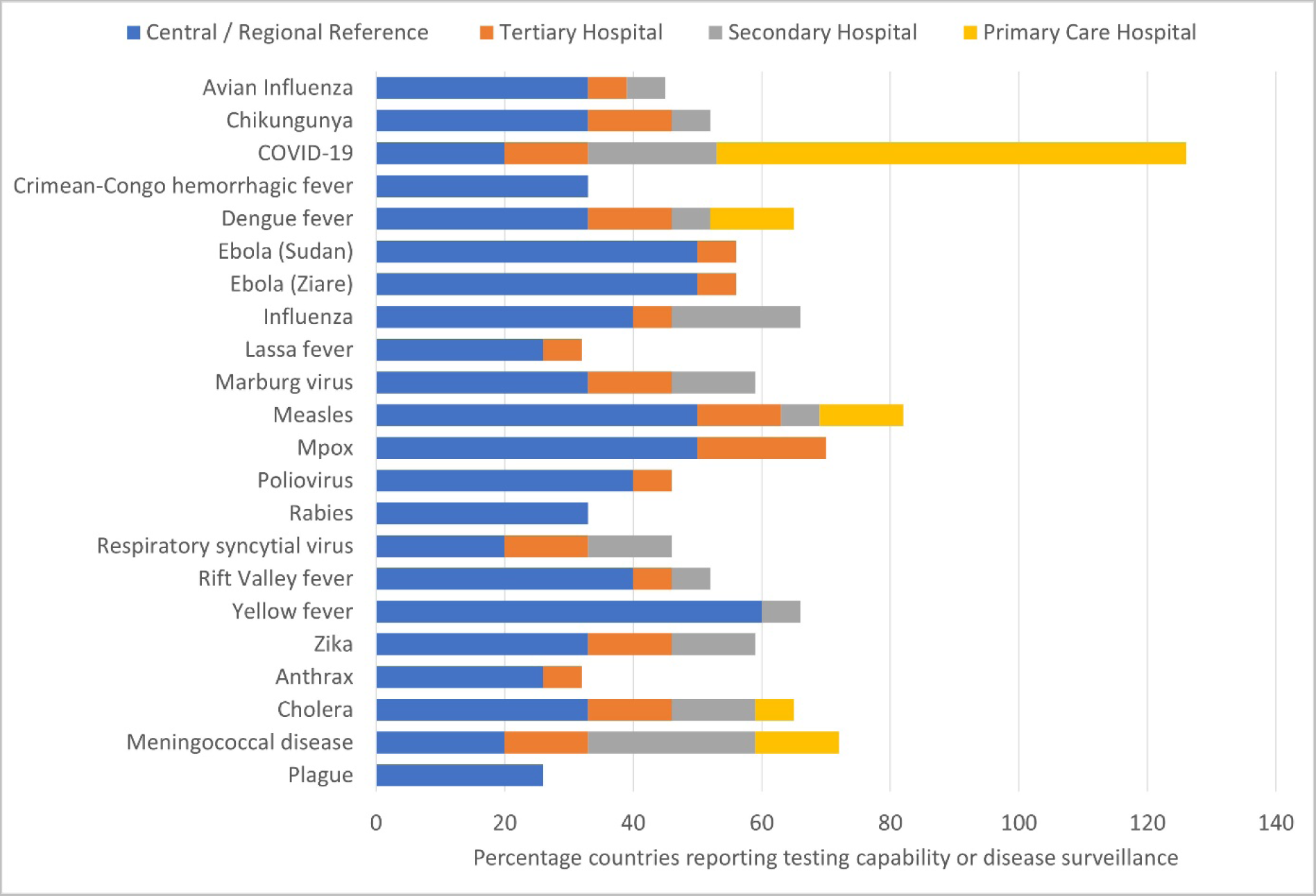
Stacked bar graph showing the percentage of countries (n=15) that can test for the specified priority epidemic-prone disease at each tier of the laboratory system.

The most common testing technology reported for diagnosing causative agents of epidemic-prone diseases was real-time polymerase chain reaction (qPCR) followed by enzyme-linked immunosorbent assay (ELISA). There were differences in testing capacity and technology across the participating countries. All 15 countries reported use of PCR testing for COVID-19. Measles and mpox were the next most common diseases diagnosed by PCR. Among Member States using PCR for testing, open-source instruments were available in 60% of national public health laboratories, 47% of regional and tertiary laboratories, and 20% of secondary care laboratories. ELISA was routinely performed to diagnose seven diseases (measles, yellow fever, CCHF, Rift Valley fever, dengue, Zika and chikungunya) across eleven countries, with measles being the most common. Among bacterial infections, few countries reported using molecular techniques for diagnosis, with between one and three countries each reporting molecular techniques for diagnosing cholera, plague, anthrax, and meningococcal disease. Two countries reported conducting culture for diagnosing cholera, plague, and meningococcal disease. Some countries did not report diagnostic capacity for testing certain infections. Eleven of the 15 countries did not test for anthrax, and ten countries did not perform testing for plague and rabies due to lack of capacity.

### Surveillance programmes for priority infectious diseases

The three most common surveillance programmes that countries were actively running during the survey were COVID-19, measles, and polio (Fig 3). Twelve countries reported having surveillance programmes for between three and twelve priority diseases. Several countries reported both the availability of PCR testing and the presence of surveillance programmes for some priority diseases, for example, plague, anthrax, cholera and meningitis. For polio, ten countries reported having surveillance programs, but only six countries indicated they have PCR diagnostic capacity.

**Fig 3.**
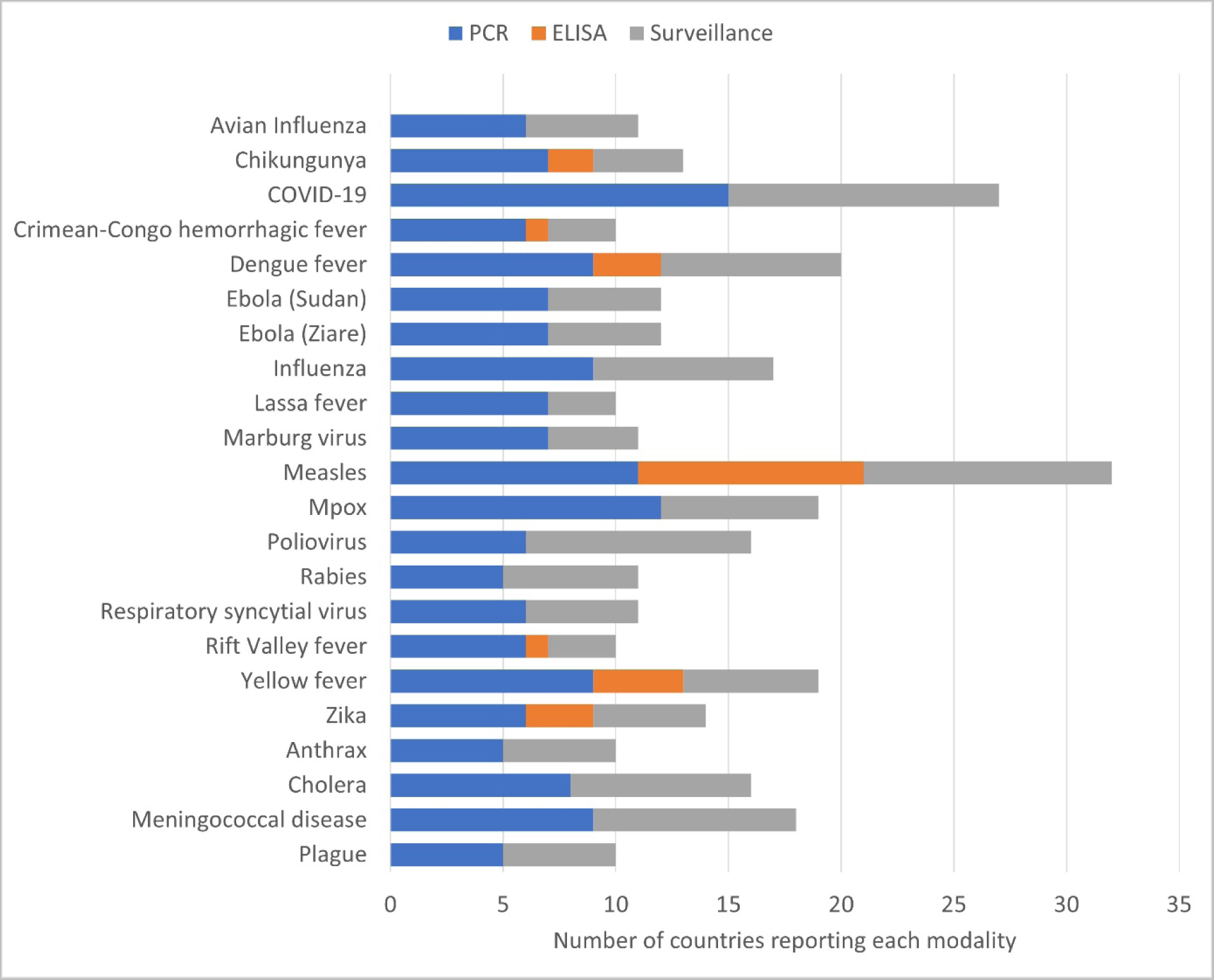
Number of countries reporting capacity for laboratory diagnosis (PCR and/or ELISA) of the priority epidemic-prone diseases and current active surveillance systems.

### Epidemic-prone disease priorities for diagnosis and surveillance

There were large differences in countries’ priorities for disease diagnosis and surveillance. Thirteen countries reported COVID-19 as the highest priority for their country, whereas two countries reported dengue fever as their highest priority. Others reported avian influenza, plague, anthrax, respiratory syncytial virus (RSV) disease, chikungunya, and EVD as their priority. There were disparities between priority and the capacity to diagnose diseases. For example, while COVID-19 was reported to be a high-priority disease with high diagnostic capacity among countries, four countries reported polio as a high-priority disease but with low diagnostic capacity using the Likert scale 1-5 for both capacity to diagnosis and priority (Fig 4).

**Fig 4.**
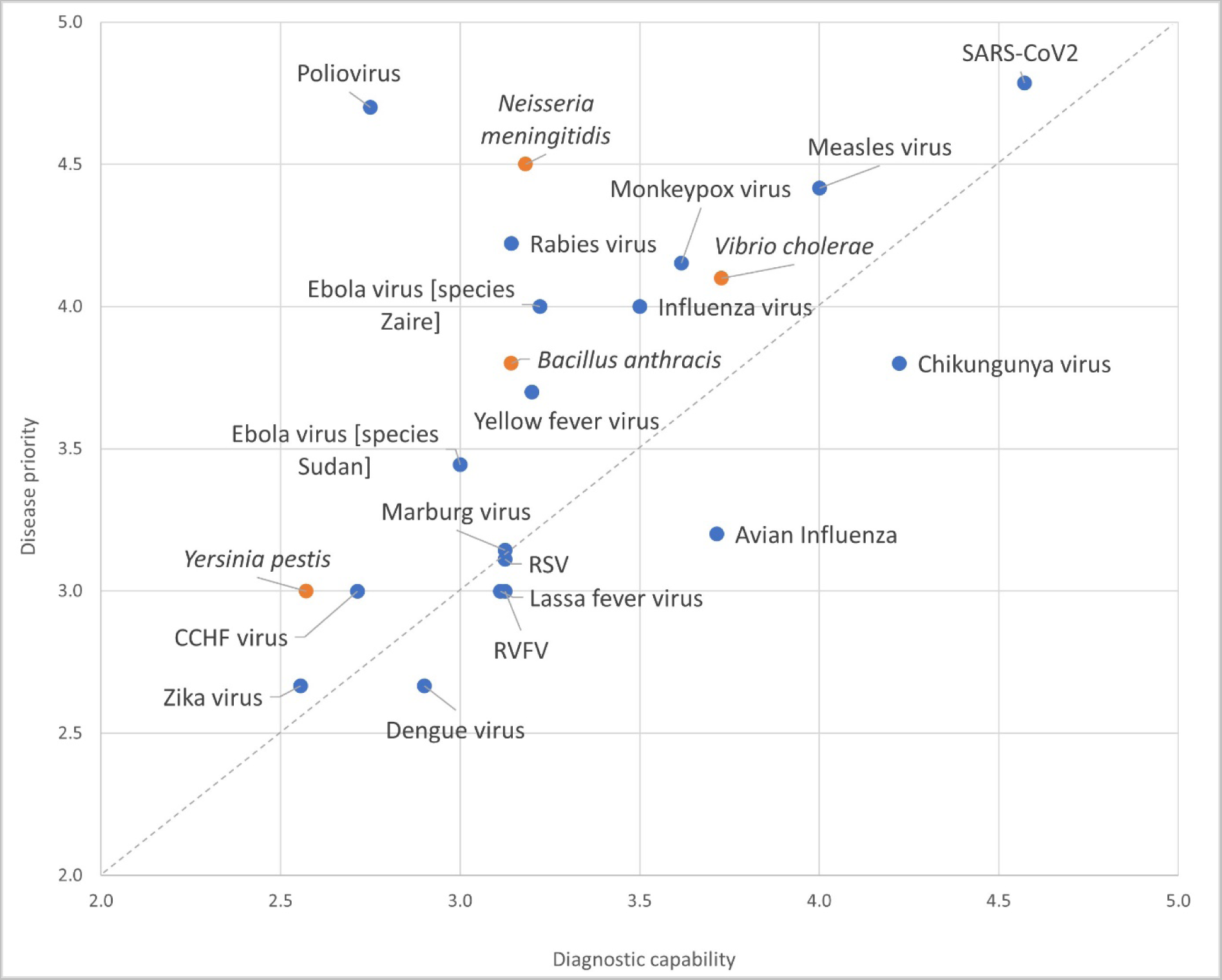
Self-reported testing capacity against the country priority for each epidemic-prone disease across the 15 Member States participating in the survey. Blue marker represents viral diseases, and the orange marker designates bacterial diseases. Inputs were provided on a Likert scale (1-5, with 5 being the highest priority), the average scores across reporting countries for the pathogen are plotted.

### Need for laboratory multiplexing testing capacity

Most respondents indicated the need for multiplex diagnostic testing capacity for infectious respiratory illnesses and arbovirus infections followed by meningitis (Table 1). For some diseases, the perceived need was less commonly reported.

**Table 1.** Number of countries reporting the need for multiplex testing by disease and/or pathogens.

| Diseases (pathogens) for which multiplex testing was reported as most needed | Number of countries<br>n=14 (%) |
| --- | --- |
| Infectious respiratory illness (influenza virus, RSV, SARS-CoV-2) | 11 (79%) |
| Arbovirus (dengue, chikungunya, Zika, yellow fever, Rift Valley fever, West Nile viruses, encephalitis) | 9 (64%) |
| Meningitis (including *Neisseria meningitidis*, *Streptococcus pneumoniae*) | 7 (50%) |
| Diarrhoeal illness including cholera | 3 (21%) |
| Viral haemorrhagic fevers (including CCHF virus, Marburg virus, Lassa virus) | 3 (21%) |

### Reported challenges limiting diagnostic capacity for future epidemics

Eighty-five per cent of countries that responded to the survey cited inconsistent laboratory supplies such as PCR reagents, extraction kits and consumables as the primary challenge to developing laboratory capacity to diagnose future epidemics. This was followed by inadequate infrastructure (45%), limited government funding (43%), inadequate equipment management (35%) and inadequate human resources (25%) (Fig 5).

**Fig 5.**
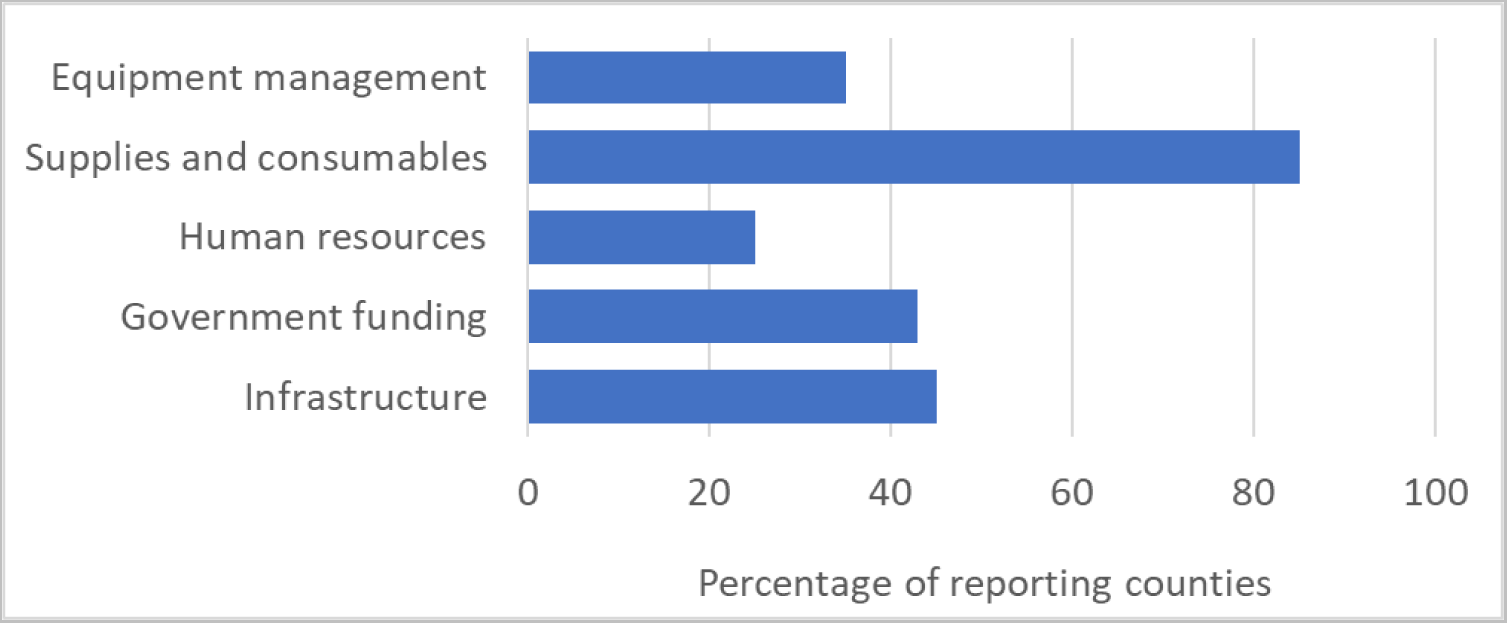
Reported gaps in laboratory diagnostics capacity limiting support for future epidemics.

## Discussion

The survey aimed to understand countries’ laboratory diagnostic needs for epidemic preparedness and response. The results offered insights into existing laboratory capacity for priority epidemic-prone diseases across 15 countries. They provided an overview of the capacity within functional tiered laboratory networks across various African Union Member States. These capacities are crucial for implementing the IHR and addressing urgent public health threats such as COVID-19, EVD, measles, CCHF, and others.

In recent years, the Africa region has experienced a surge in outbreaks and epidemics, threatening the health of populations and socio-economic stability, with potential global ramifications (12). Reported disease outbreaks include yellow fever, Rift Valley fever, mpox, measles, CCHF, dengue fever, chikungunya, EVD, Marburg virus disease, cholera, diphtheria, and COVID-19. However, Africa’s inadequate disease diagnosis and surveillance systems hinder the prediction, prevention and effective management of emerging infectious diseases (13). Delayed detection of outbreaks is attributed to a lack of real-time surveillance, suboptimal laboratory networks, limited diagnostic capabilities, inadequate resources, and insufficient technical and managerial capacities.

This survey showed that the capacity for diagnosing infectious diseases varied by disease, country, and laboratory level within tiered networks. The most common technology for diagnosing epidemic-prone diseases was PCR, followed by ELISA. All 15 participating countries reported the use of PCR testing for COVID-19, highlighting the improvements in diagnostics from the COVID-19 pandemic. In contrast, priority diseases such as polio and plague had limited PCR testing capacity. These results may reflect differing country priorities, levels of laboratory testing maturity, and/or the availability of national and external funds to diagnose priority pathogens. The findings suggest that policy and decision-makers have shown a consistent commitment to improving laboratory capacity and system development, as demonstrated by the high capacity to diagnose COVID-19; however, our assessment has also uncovered significant deficiencies in disease diagnostic capacity, particularly at different levels of the tiered laboratory networks that handle epidemic-prone diseases.

Lower-level laboratories exhibited weaknesses in testing for diseases such as Rift Valley fever, CCHF, plague, dengue fever and Zika, which were predominantly conducted at central and regional laboratories, compared to their capabilities for diagnosing COVID-19, polio, measles, mpox, cholera and meningitis. This disparity reflects historical prioritisation and investment in testing for specific diseases, technological advancements, biosafety concerns, and the diversity and availability of funding resources. The COVID-19 crisis underscores the world’s historical unpreparedness to detect and respond to emerging infectious diseases (14). Recent outbreaks of diseases such as Lassa fever, EVD, Marburg virus disease, diphtheria, anthrax, and cholera across the continent highlight the urgent need to strengthen diagnostic capacity for emergency response and outbreak management. Lassa fever’s expanding geographical spread in West Africa underscores the evolving epidemiology of infectious diseases and the necessity for all countries to possess resilient and adaptable diagnostic capabilities (15).

Inadequate diagnostic testing remains a significant issue in African countries (16, 17), particularly at lower-level laboratories. Many commercially available diagnostic tools do not meet the needs of these laboratories or cannot be used due to weak infrastructure. Increased investment in testing capacity at lower-level laboratories is crucial, along with developing and implementing appropriate technologies for diagnosing epidemic-prone diseases. Some tests are available in multiplex format, enabling the detection of multiple pathogens in a single sample for both surveillance and clinical management purposes. Establishing diagnostic capacity for priority diseases of epidemic potential through integrated testing within existing laboratory capacity and with multiplex testing will improve epidemic preparedness, aid early detection and response, and help prevent outbreaks from becoming pandemics (18, 19).

Limited access to diagnostic testing has been a bottleneck in the early detection of priority diseases in Africa (20–22). Diagnostic capacity has often focused on diseases such as HIV, TB, malaria, and more recently, COVID-19 (23, 24). However, laboratories and health systems established for these infections can also be used to detect other priority diseases of epidemic potential, thereby expanding the range of pathogen detection capacity in African Union Member States using existing methodologies.

This survey highlights that the effectiveness of response measures is impeded by various factors, including limited laboratory capacities, difficulties in accessing diagnostic services, shortages of human resources, and fragmented surveillance systems. These challenges exacerbate the complexities of response efforts in numerous countries within the Africa region. The integration of testing and effective use of existing diagnostic platforms and laboratory infrastructure were demonstrated with SARS-CoV-2 testing during the COVID-19 pandemic.

The limitations of this survey included reliance on self-reporting and the limited number of countries that responded. Despite these limitations, the survey results provided valuable insights into diagnostic capacity, disease priorities, surveillance systems and current challenges to the diagnosis of epidemic-prone diseases and has highlighted gaps that urgently need to be addressed to enhance epidemic preparedness, facilitate early disease detection and response, and mitigate the risk of outbreaks escalating into pandemics.

## Conclusion

The increased technical capacity for diagnosing infectious diseases since the COVID-19 epidemic is evident. With ongoing outbreaks of diseases such as yellow fever, Rift Valley fever, mpox, measles, CCHF, dengue fever, chikungunya, EVD, Marburg virus disease, cholera and diphtheria across the continent, there is an urgent need to address the gaps in diagnostic capacity and disease surveillance. Recognising the varying country priorities, expanding point-of-care and multiplex testing for syndromic disease surveillance, and monitoring changes in epidemiology are crucial steps to enhance readiness for detecting epidemic-prone diseases. Countries need to prioritise rapid diagnostic tests and portable technologies while strengthening national and regional laboratory networks. This requires updating infrastructure, improving the laboratory supply chain, and investing in staff training and development.

## Data Availability

The data that support the findings of this manuscript are readily accessible without any limitations

## Acknowledgements

We express our gratitude to the Ministry of Health of participated countries and to the numerous colleagues at our respective institutions who have engaged in discussions, provided valuable input, and offered feedback on the paper. Their contributions have significantly enhanced the quality of the final manuscript. The authors would like to express their gratitude to Africa Laboratory Technical Working Group (AfLTWG) members. The contents of this paper are solely the responsibility of the authors.

## Supporting information

S1 laboratory capacity questionnaire (XLS) S2 list of epidemic-prone diseases (DOCX)

